# Cost and availability of drugs and treatment regimens and availability of drug resistance testing for tuberculosis in Europe

**DOI:** 10.1101/2022.02.15.22271006

**Authors:** Gunar Günther, Lorenzo Guglielmetti, Claude Leu, Christoph Lange, Frank van Leth, TBnet

## Abstract

**Background:** Access to comprehensive diagnostics and novel anti-tuberculosis medicines is crucial to improve tuberculosis control at times of emerging *Mycobacterium tuberculosis* drug resistance.

**Methods:** We investigated access to genotypic and phenotypic *M tuberculosis* drug susceptibility testing (DST), availability of anti-tuberculosis drugs and calculated cost of drugs and treatment regimens at major tuberculosis treatment centers in countries of the World Health Organization (WHO) European region. Results are stratified by middle and high-income countries.

**Results:** Overall, 43 treatment centers in 43 countries participated in the study. Phenotypic DST was available for WHO group A drugs levofloxacin/moxifloxacin, bedaquiline and linezolid, in 75%/82%, 48%, and 72% of countries, respectively. Overall, 84% and 56% of countries had access to bedaquiline and delamanid, while only 14% had access to rifapentine. Median cost of regimens for drug-susceptible tuberculosis, multidrug-resistant tuberculosis (shorter regimen, including bedaquiline for six months) and pre-extensively drug-resistant tuberculosis (including delamanid) were €44, €764 and €7 094 in middle income countries (n=12), and €280, €29 765, €207 035 in high income countries (n=29).

**Conclusion:** Tuberculosis control in Europe is limited by widespread lack of DST capacity to new and re-purposed drugs, lack of access to essential medications and high treatment cost for drug-resistant tuberculosis.

**Research in context:** *Evidence before this study:* Availability and access to anti-tuberculous treatment are essential for optimal treatment outcomes. Newly developed drugs like bedaquiline have demonstrated an enormous potential to improve outcomes, in particular for the treatment of drug-resistant tuberculosis. However, data on availability and cost of tuberculosis drugs and regimens are scarce. We searched PubMed for original research that reported the cost of anti-tuberculosis drugs and regimens across multiple countries in the WHO European region since Jan 1, 2012. The Pubmed search ((cost[MeSH Major Topic]) AND (tuberculosis[MeSH Major Topic]) AND [(treatment[MeSH Major Topic]) OR (drug[MeSH Major Topic])] AND (“2012/01/01”[Date -Entry] : “3000”[Date - Entry])) did not reveal any comprehensive data on cost of anti-tuberculosis drugs since the introduction of new (bedaquiline, delamanid, pretomanid) and re-purposed drugs in the WHO European region. Recent information on availability of ***Mycobacterium tuberculosis*** drug susceptibility testing is limited to a single, laboratory-based survey.

*Added value of this study:* Building on a previous study, performed by the Tuberculosis Network European Trialsgroup (TBnet) in 2013, the current study documents a) the concerning lack of diagnostic capacity of drug susceptibility testing for new and repurposed anti-tuberculosis drugs; b) the lack of availability of adequate regimens for the treatment of multidrug-resistant and (pre-) extensively drug-resistant tuberculosis, in particular in middle income countries; and c) the enormous cost of regimens for the treatment of drug-resistant tuberculosis, in particular in high-income countries.

*Implications of all the available evidence:* The lack of availability of drug-resistance testing in the presence of new and re-purposed drugs bears the high risk of undetected amplification of *M tuberculosis* drug resistance. In addition, it implies that identification of patients with extensively drug-resistant tuberculosis is currently not possible in many countries in the WHO European region. The cost of drugs and regimens for drug-resistant tuberculosis treatment are very high compared to those for drug-susceptible tuberculosis and highly variable across different countries. Access to adequate treatment regimens for (pre-) extensively drug-resistant tuberculosis is suboptimal. Rapid upscaling of comprehensive *M tuberculosis* drug resistance testing and provision of novel anti-tuberculosis drugs are urgently required to provide patients affected by drug-resistant tuberculosis with adequate treatment regimens and prevent the emergence of additional drug resistance in *M tuberculosis* naturally occurring under insufficient treatments.

## Introduction

Diagnostic improvements and availability of novel anti-tuberculosis medicines have brought substantial change to the management of patients affected by drug-resistant tuberculosis.^1,2^ Molecular drug susceptibility testing (DST) based on nucleic acid amplification technologies entered clinical routine in many countries.^3^ New anti-tuberculosis drugs (i.e. bedaquiline, delamanid and pretomanid) were approved for drug-resistant tuberculosis treatment along with fundamental changes in treatment guidelines and regimens.^4^ Although with relevant delays, phenotypic DST for new and re-purposed drugs (i.e. linezolid, clofazimine) are being developed.^3^ Such innovations can improve tuberculosis control if they are accessible for patients and programs. Improving access by ensuring affordable pricing of drugs is a crucial issue for all new anti-tuberculosis drugs, and a major topic of political debate and advocacy.^5^

Following a recent revision of the hierarchy of anti-tuberculosis drugs for the treatment of patients with drug-resistant tuberculosis by the World Health Organization (WHO) in 2020,^6^ little is known about the availability of drugs and DST for new and re-purposed anti-tuberculosis drugs. The same holds true for cost of these drugs and treatment regimens.^7,8^ The Tuberculosis Network European Trialsgroup (TBnet), an European-based network promoting TB research and training,^9^ first evaluated the availability and cost of anti-tuberculosis drugs and regimens among 37 European countries in 2013,^10^ at a time when bedaquiline, delamanid and pretomanid were not yet available. In order to provide an updated account on the availability of anti-tuberculosis DST and the costs and availability of anti-tuberculosis drugs and regimen, we performed a new survey among major treatment centres and through TBnet referent physicians in the countries of the WHO European region.

## Methods

### Data collection

Data on tuberculosis drug availability, cost, and availability of DST for all anti-tuberculosis drugs were surveyed administering a standardized questionnaire to TBnet representatives with experience in the management of drug-resistant tuberculosis at referral treatment centres in countries of the WHO European region. If no TBnet representatives were available in a country, we searched Pubmed for major publications on drug-resistant tuberculosis and approached respective authors from target countries. Data collection for cost and DST availability was performed from June to December 2020 and updated in October 2021. The list of the drugs in the survey was developed with reference to those available via the global drug facility.^11^

### Data analysis

Drug cost calculations were based on available formulations and cost for one unit (tablet or vial) of the drug. We determined the number of units required to provide adequate daily treatment for patients with 70 kg of body weight, according to WHO-recommended drug doses.^4^ When available, fixed dose combinations (FDC) were included in the calculation of the regimen cost and the least-expensive regimen option was reported. Daily treatment cost for drugs given on a non-daily basis, like bedaquiline, were based on weekly cost divided by seven. Cost data were collected in local currency. If data were given in US dollars (USD), the exchange rate as of 01.07.2020 was used to convert in local currency. Costs are reported in Euro (€) when there is no direct between-country comparison. For direct between-country comparisons, drug cost were converted in international dollars (ID$) using the purchasing power parity conversion factor from the international comparison program 2017.^12^ Stratification according to income followed the World Bank classification, where upper and lower middle income countries are combined as middle income countries (figure S1).^13^ Costs of regimens and drugs are presented as median with minimum and maximum values, if not otherwise stated.

We selected regimens for drug-susceptible tuberculosis (DS-TB), multidrug-resistant/rifampicin-resistant tuberculosis (MDR/RR-TB), pre-extensively drug-resistant tuberculosis (pre-XDR TB), and extensively drug-resistant tuberculosis (XDR-TB) based on latest guidelines from WHO^4,14^, and American Thoracic Society/Centers for Disease Control/European Respiratory Society/Infectious Diseases Society of America (ATS/CDC/ERS/IDSA)^15^; regimen compositions are shown in table S1. DS-TB was defined as susceptible to all first-line TB drugs. MDR/RR-TB, pre-XDR TB, and XDR-TB were defined according to WHO 2020 definitions.^16^ We hereby present results for eight priority regimens (table 1). Results for additional regimens are available in the supplement (table S2). We did not present cost for a standardized regimen containing bedaquiline, linezolid and pretomanid (BPaL)^17^, as the regimen is currently only recommended by WHO in the context of operational research. DST availability was evaluated for the same list of drugs as cost data and stratified by phenotypic and genotypic testing. In the questionnaire, clinicians were not asked to specify whether DST was performed locally at the tuberculosis treatment centre.

**Table 1:**
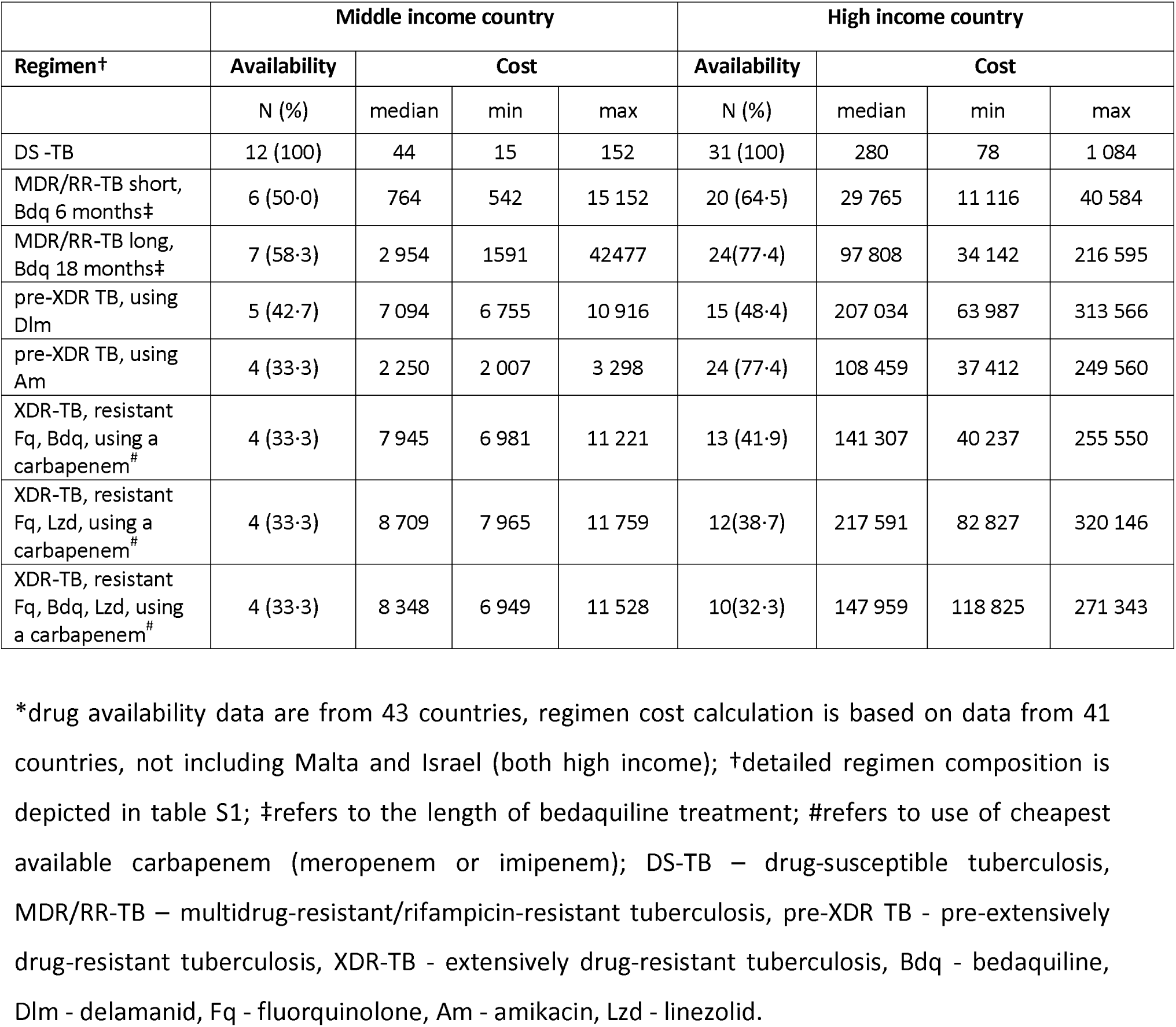
Availability and cost of drug regimens for the treatment of tuberculosis in countries in the WHO Europe region, stratified by world bank income classification*, in Euros.

|  | Middle income country |  |  |  | High income country |  |  |  |
| --- | --- | --- | --- | --- | --- | --- | --- | --- |
| Regimen† | Availability | Cost |  |  | Availability | Cost |  |  |
|  | N (%) | median | min | max | N (%) | median | min | max |
| DS -TB | 12 (100) | 44 | 15 | 152 | 31 (100) | 280 | 78 | 1 084 |
| MDR/RR-TB short, Bdq 6 months‡ | 6 (50·0) | 764 | 542 | 15 152 | 20 (64·5) | 29 765 | 11 116 | 40 584 |
| MDR/RR-TB long, Bdq 18 months‡ | 7 (58·3) | 2 954 | 1591 | 42477 | 24(77·4) | 97 808 | 34 142 | 216 595 |
| pre-XDR TB, using Dlm | 5 (42·7) | 7 094 | 6 755 | 10 916 | 15 (48·4) | 207 034 | 63 987 | 313 566 |
| pre-XDR TB, using Am | 4 (33·3) | 2 250 | 2 007 | 3 298 | 24 (77·4) | 108 459 | 37 412 | 249 560 |
| XDR-TB, resistant Fq, Bdq, using a carbapenem# | 4 (33·3) | 7 945 | 6 981 | 11 221 | 13 (41·9) | 141 307 | 40 237 | 255 550 |
| XDR-TB, resistant Fq, Lzd, using a carbapenem# | 4 (33·3) | 8 709 | 7 965 | 11 759 | 12(38·7) | 217 591 | 82 827 | 320 146 |
| XDR-TB, resistant Fq, Bdq, Lzd, using a carbapenem# | 4 (33·3) | 8 348 | 6 949 | 11 528 | 10(32·3) | 147 959 | 118 825 | 271 343 |

\*drug availability data are from 43 countries, regimen cost calculation is based on data from 41 countries, not including Malta and Israel (both high income); †detailed regimen composition is depicted in table S1; ‡refers to the length of bedaquiline treatment; #refers to use of cheapest available carbapenem (meropenem or imipenem); DS-TB – drug-susceptible tuberculosis, MDR/RR-TB – multidrug-resistant/rifampicin-resistant tuberculosis, pre-XDR TB - pre-extensively drug-resistant tuberculosis, XDR-TB - extensively drug-resistant tuberculosis, Bdq - bedaquiline, Dlm - delamanid, Fq - fluorquinolone, Am - amikacin, Lzd - linezolid.

### Ethics

Ethical clearance was granted by the Institutional Review Board of Bligny Hospital, France (January 15th, 2020; CRE 2020 01). As no patient data were collected, ethical board review was not applicable at any of the participating centres.

## Results

### Survey response

The WHO European region has 53 countries (not including Kosovo). We excluded Central Asian countries and small city countries (in total, n=8) from the survey and therefore did not contact representatives from Andorra, Kazakhstan, Kyrgyzstan, Monaco, San Marino, Tajikistan, Turkmenistan, and Uzbekistan. Overall, data on drug availability were obtained and analysed from 43, data on drug cost from 41, and data on DST availability from 40 countries. We were unable to obtain responses on drug cost, availability, and DST availability from Azerbaijan, Bosnia and Herzegovina, and Montenegro. Drug cost data were not available from Malta and Israel. DST data were not available from Malta, Kosovo and Iceland.

### Availability of DST

Phenotypic DST testing was generally more widely available than genotypic testing. While phenotypic DST for all first-line drugs was available in 38/40 (95%) countries, genotypic DST was available for rifampicin in 40/40 (100%) and for isoniazid in 38/40 (95%) countries, but only in 21/40 (53%) for ethambutol and in 12/40 (30%) for pyrazinamide. For WHO Group A drugs, the frequency of countries with availability of phenotypic and/or genotypic DST was as follows: 30/40 (75%) and 29/40 (73%) for levofloxacin, 33/40 (82%) and 32/40 (80%) for moxifloxacin, 19/40 (48%) and 10/40 (25%) for bedaquiline, 29/40 (72%) and 11/40 (28%) for linezolid, respectively (figure 1). For Group B drugs, the frequency of countries with availability of phenotypic and/or genotypic DST was 25/40 (63%) and 11/40 (28%) for clofazimine, and 23/40 (58%) and 8/40 (20%) for cycloserine/terizidone, respectively. Among Group C drugs, phenotypic and/or genotypic DST testing was only available in 6/40 (15%) and 1/40 (2.5%) countries for carbapenems (meropenem and imipenem), and in 17/40 (42%) and 10/40 (25%) for delamanid, respectively. Phenotypic DST for rifapentine could not be evaluated in any of the countries and genotypic DST for this drug was only available in 6/40 (15%) countries. Similarly, phenotypic DST to pretomanid was only available in 2/40 (5%) and genotypic DST in 4/40 (10%) of the countries, respectively.

**Figure 1:**
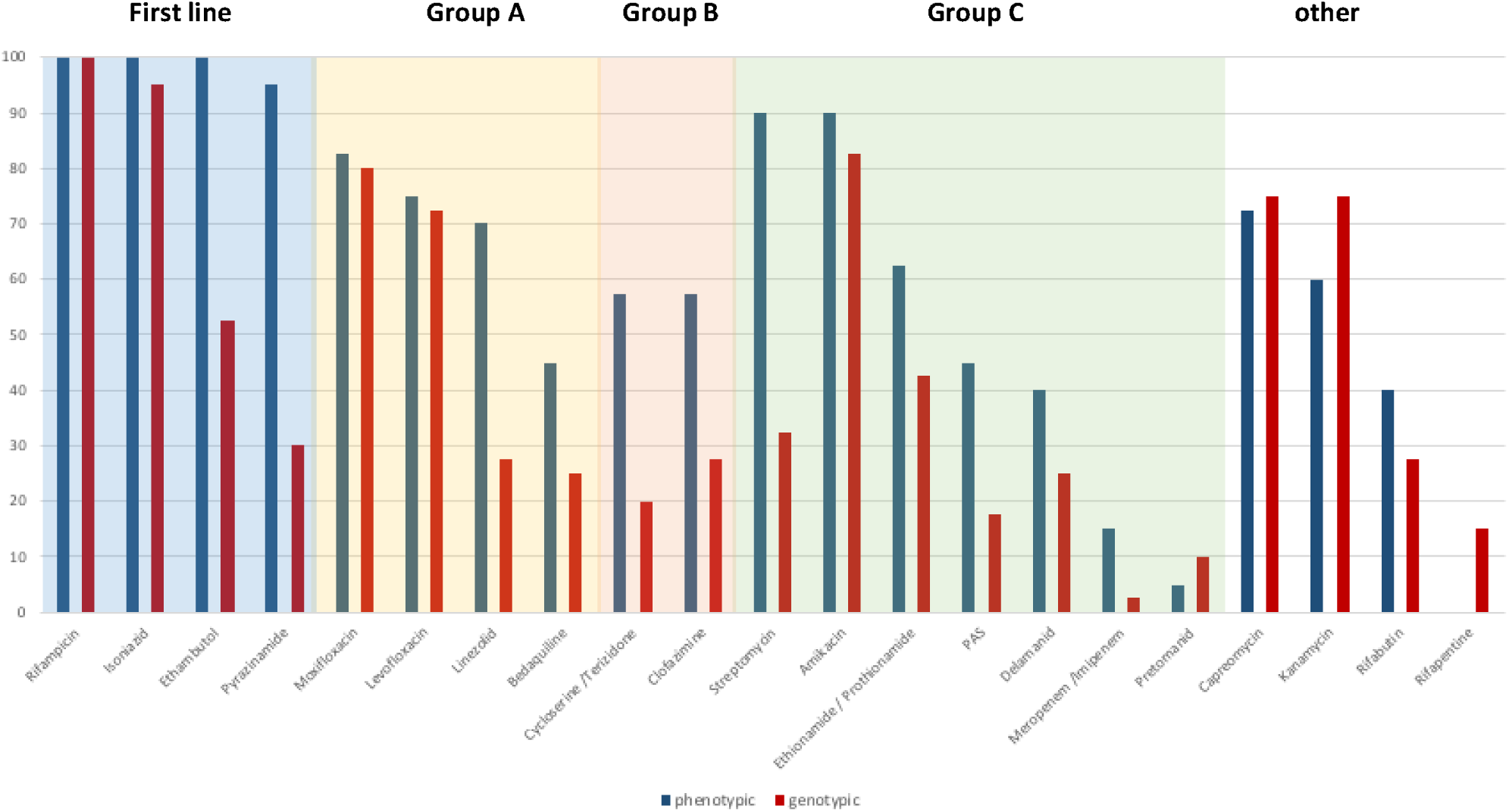
Availability of phenotypic and genotypic drug susceptibility testing to tuberculosis drugs in countries in the WHO European region*, in %. *n=40 countries; Kosovo, Iceland and Israel did not provide data on availability of drug susceptibility testing

**Figure 2:**
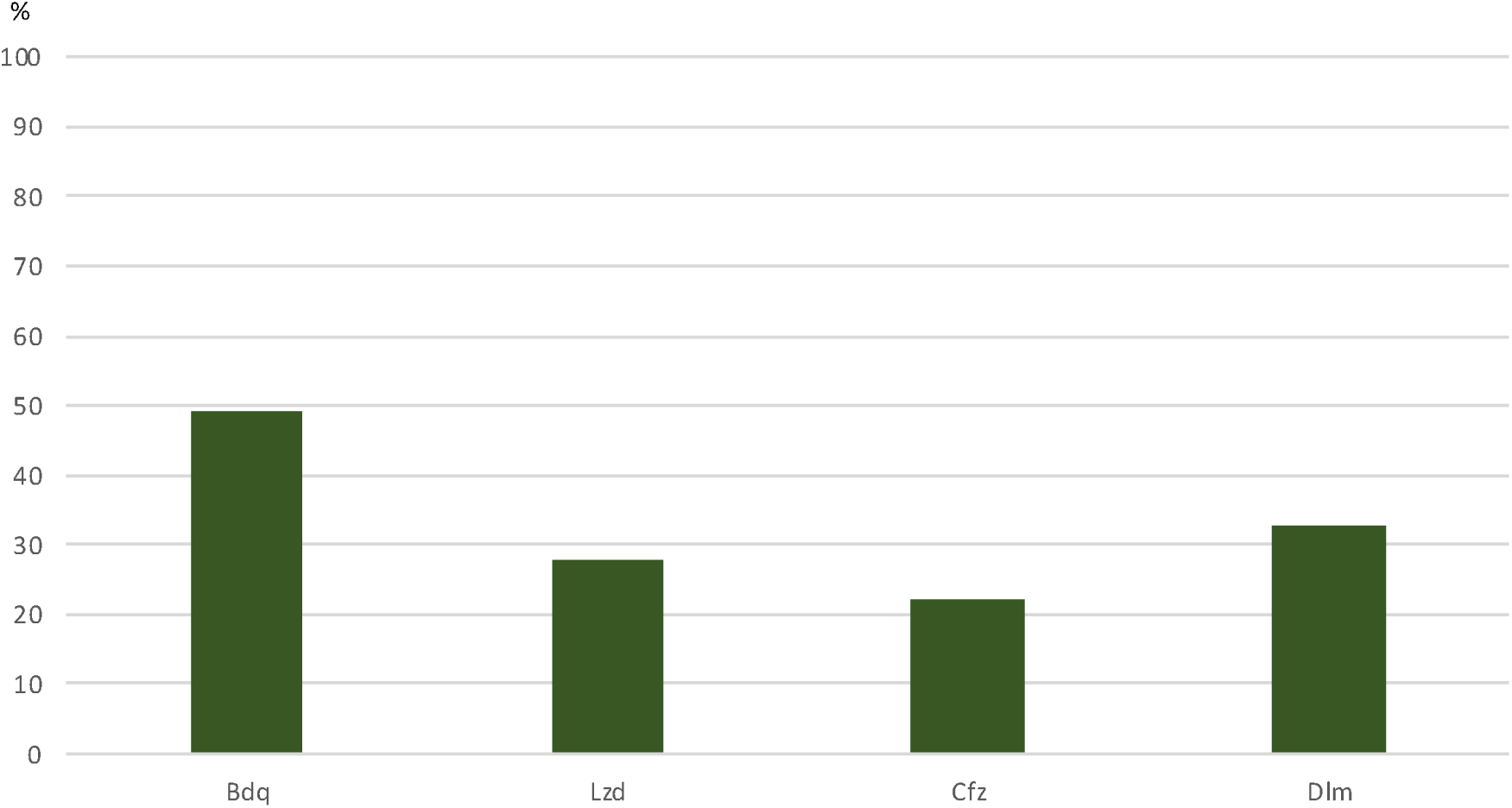
Proportion of countries with availability of bedaquline, linezolid, clofazimine, and delamanid, but absence of drug susceptibility testing for those drugs*. *n=40 countries; Kosovo, Iceland and Israel did not provide data on availability of drug susceptibility testing Bdq - bedaquiline, Lzd – linezolid, Cfz – clofazimine, Dlm – delamanid,

### Availability of tuberculosis drugs

The four first-line drugs rifampicin, isoniazid, pyrazinamide and ethambutol were available in all 43 countries, as single drugs or as part of fixed-dose drug combinations. Levofloxacin and moxifloxacin were available in 43/43 (100%) and 41/43 (95%), bedaquiline in 36/43 (84%) and linezolid in 43/43 (100%) countries, respectively. Clofazimine was available in 35/43 (81%) countries, but only in 8/12 (67%) middle income countries. Delamanid was available in 24/43 (56%) countries. Meropenem and imipenem were available in 28/43 (65%) and 25/43 (58%) countries: access was in particular limited in middle income countries, where only 3/12 (25%) and 4/12 (33%) countries had access to the drugs, compared to 25/31 (81%) and 21/31 (68%) high income countries. Pretomanid was available only in 4/43 (9%) countries, all high income countries. Only 6/43 (14%) countries reported access to rifapentine (table S4).

### Cost of tuberculosis drugs

Tuberculosis drugs were generally less expensive in middle income countries, albeit there was large variability in drug cost between countries. The drugs with the highest median daily treatment costs were delamanid, bedaquiline, and rifapentine in high income countries and imipenem, meropenem, and delamanid in middle income countries, respectively. Daily median treatment costs for delamanid were €128·04 in high income countries, and €8·52 in middle income countries, while for bedaquiline it was €103·98 and €1·60, respectively. The daily median treatment cost of amikacin was €10·10 in high income countries and €1·18 in middle income countries (table S4).

### Availability of tuberculosis treatment regimens

Treatment of DS-TB according to current WHO guidelines was available in all 43 (100%) countries (table 1). The shorter MDR/RR-TB regimen with bedaquiline was available in 26/43 (60%) countries, while the conventional long MDR/RR-TB regimen was available in 31/43 (72%) countries. A pre-XDR TB treatment, with amikacin or delamanid replacing the fluoroquinolones, was available in 28/43 (65%) and 20/43 (47%) countries, respectively. Treatment of patients with XDR-TB with a regimen including a carbapenem was only available in 17/43 (40%) countries (tables 1 and S2).

### Cost of tuberculosis treatment regimens

In general, regimens were considerably less expensive in middle income than in high income countries. Figure 3 illustrates the direct comparison of the cost of treatment regimens between countries, taking into account purchasing power parity based in ID$. Figure 4 shows the overall distribution of regimen costs, based on Euro. The median cost of a DS-TB regimen was €44 in middle income countries and €280 in high income countries. The median cost of the shorter MDR/RR-TB regimen with bedaquiline for 6 months was €764 in middle income countries and €29 765 in high income countries, while the conventional long MDR/RR-TB treatment regimen with bedaquiline for 6 months costed €2 214 and €51 617, respectively (tables 1, S2 and S3). A pre-XDR TB treatment regimen using delamanid or amikacin costed €7 094 or €2 250 in middle income countries, respectively, and €207 034 or €108 459 in high income countries, respectively. A regimen for the treatment of patients with XDR-TB with resistance to fluoroquinolones and linezolid, including bedaquiline, delamanid, and a carbapenem costed €8 709 in middle income countries and €217 591 in high income countries.

**Figure 3:**
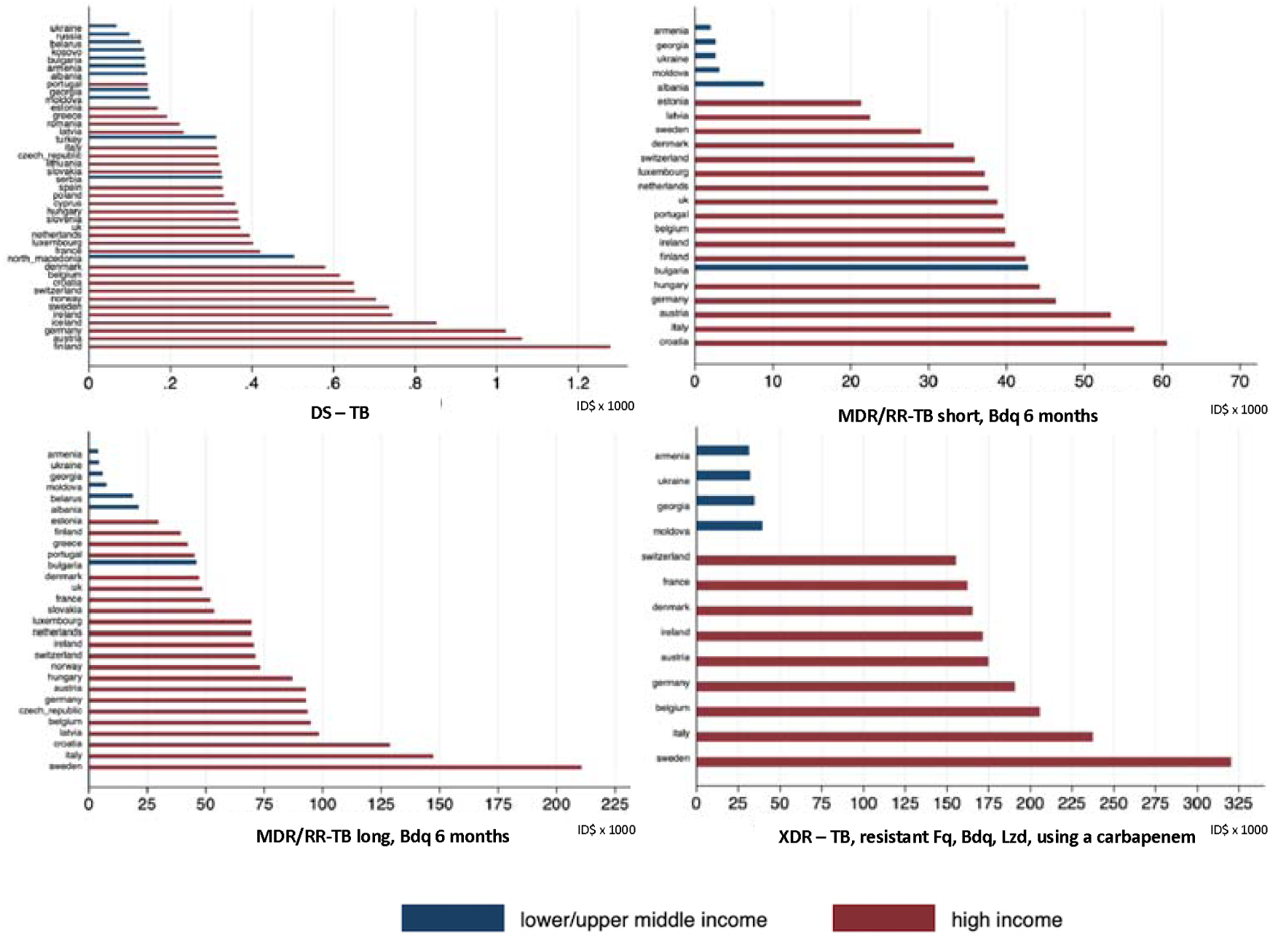
Cost of regimens for drug-susceptible tuberculosis, multidrug-resistant/rifampicin-resistant tuberculosis using the shorter regimen for nine months with six months of bedaquiline, multidrug-resistant/rifampicin-resistant tuberculosis with the long conventional regimen with six months of bedaquiline, and extensively drug-resistant tuberculosis with resistance to fluoroquinolones, linezolid, and bedaquiline, stratified by country and income classification of world bank*, in international dollars (ID$). * n=41 countries, Malta and Israel (both high income) did not provide data on drug cost; detailed regimen composition in table S1; DS-TB – drug susceptible TB, MDR/RR - multidrug-resistant/rifampicin-resistant tuberculosis, XDR - extensively drug-resistant tuberculosis, Fq – fluoroquinolones, Bdq - bedaquiline, Lzd – linezolid.

**Figure 4:**
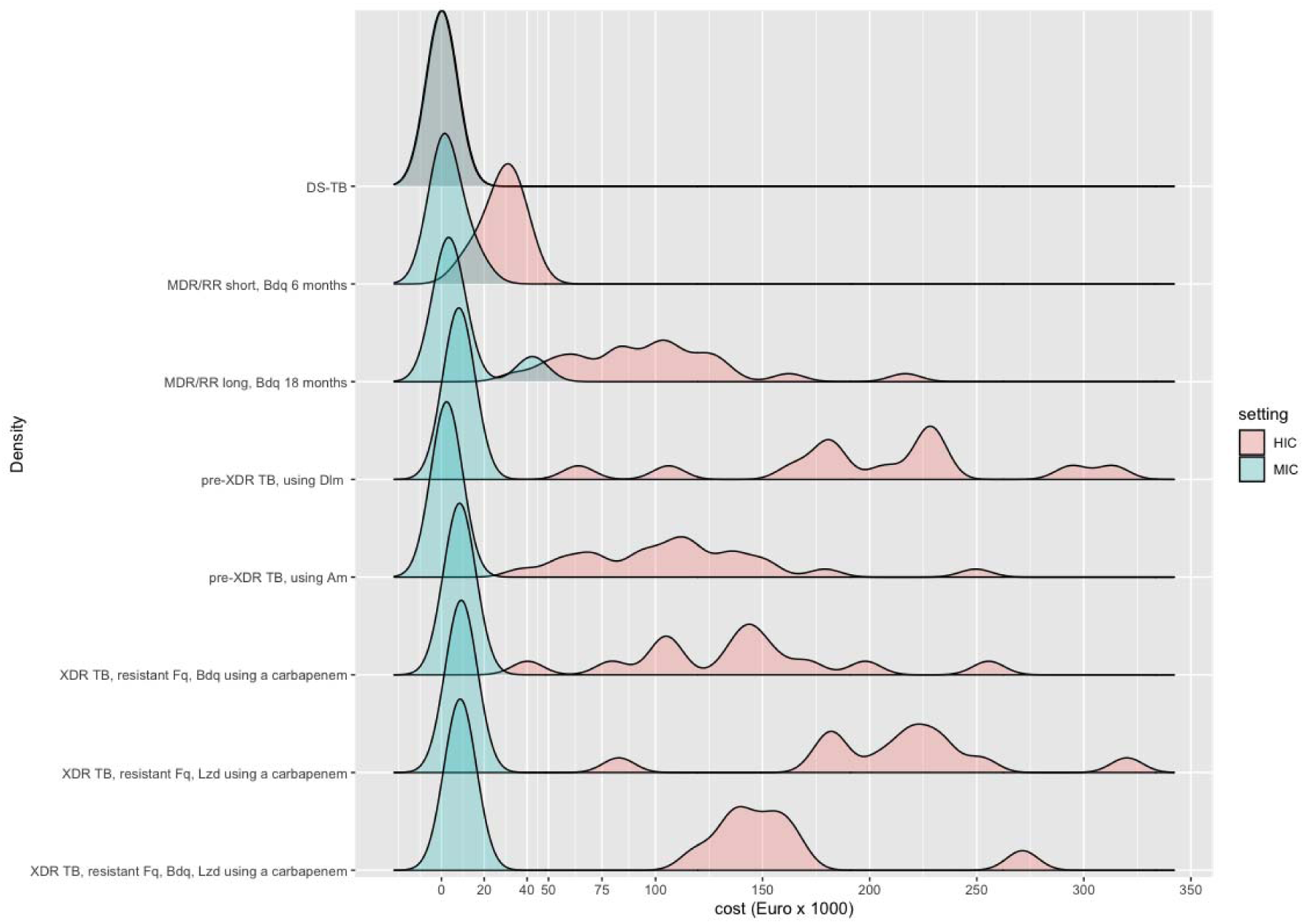
Density graph of distribution* of cost for tuberculosis drug regimens in the WHO Europe region, according to resistance status and world bank income classification (high income countries in red, middle income countries in green), in Euros. * The density graph illustrates the distribution of the cost within a given resistance pattern. The area under the curve is scaled to one (1). The height in the distribution shows the range of the cost for the majority of countries. The width of the graph shows the range of the costs observed. n=41 countries, Malta and Israel (both high income) did not provide data on drug cost HIC – high income countries, MIC – middle income countries, DS-TB - drug susceptible TB, MDR/RR-TB – multidrug-resistant/rifampicin-resistant tuberculosis, pre-XDR TB – pre-extensively drug-resistant tuberculosis, XDR-TB – extensively drug-resistant tuberculosis, Bdq - bedaquiline, Fq - fluoroquinolones, Lzd - linezolid, Dlm – delamanid, Am – amikacin.

Overall, high costs for drug-resistant tuberculosis treatment regimens were driven by specific drugs, such as delamanid and carbapenems in middle income countries, and delamanid, bedaquiline, and to a lesser extent, para-aminosalycilic acid and cycloserine/terizidone in high income countries (table S4).

## Discussion

Treatment of drug-resistant tuberculosis has undergone major changes recently due to diagnostic and therapeutic breakthroughs, including the use of new and re-purposed drugs.^4,15^ Following a first survey in 2013, we provide an updated report on the availability of DST for anti-tuberculosis drugs, as well as the costs and availability of anti-tuberculosis drugs in the WHO European region. The main finding of this survey is that availability of DST for second-line anti-tuberculosis drugs, in particular new and re-purposed drugs, is severely limited in Europe and that new drugs are more frequently available than their specific DST. Cost of drugs and regimens for drug-resistant tuberculosis treatment are very high compared to treatment of DS-TB. In addition, the cost of regimens is highly variable across different countries. Access to adequate treatment regimens for pre-XDR and XDR-TB is limited, in particular in middle income countries. Finally, almost no country in Europe has access to drugs included in new promising regimens for drug-susceptible and drug-resistant tuberculosis, such as rifapentine and pretomanid.

Phenotypic DST remains the gold standard for the detection of drug resistance in *M tuberculosis*. International guidelines for the management of drug-resistant tuberculosis suggest including in a treatment regimen at least four effective drugs, ideally based on DST results.^4,15^ Our findings are very worrying: in 2021, only 75% and 82% of the countries in the survey report phenotypic DST capacity for levofloxacin and moxifloxacin, 48% for bedaquiline, and 72% for linezolid, while availability of these drugs is much higher. Hence, physicians in many countries of the WHO European region use Group A drugs for MDR/RR-TB treatment without capacity to detect resistance against these drugs. Similar findings were obtained by the network of European tuberculosis reference laboratories.^18^ Phenotypic DST for Group B drugs clofazimine and cycloserine/terizidone is available in less than 65% of the countries, while only 42% of countries can test for delamanid resistance. In addition to the lack of DST capacity, reliable data on clinical breakpoints, compounds for resistance testing and adequate strains for external quality control are still missing for some drugs, in particular for the new drugs bedaquiline, delamanid and pretomanid.^19^ Recent reports of growing resistance to new and re-purposed drugs underline the need for resistance detection and surveillance.^20-22^ This is particularly important since resistance to Group A drugs in particular is associated with worse MDR/RR-TB treatment outcomes.^23,24^

Globally, the implementation of the new WHO definition for XDR-TB is problematic.^25,26^ The definition of XDR-TB requires confirmed resistance for rifampicin, isoniazid, a fluoroquinolone and at least another group A drug, linezolid and/or bedaquiline.^16^ According to our results, 52% of European countries cannot detect bedaquiline resistance and 27% cannot detect linezolid resistance: as a consequence, the identification of patients with XDR-TB is not possible in the majority of countries in the region. As for other processes in evolution, the main drivers of antimicrobial drug-resistance development are variation and selection. In order to avoid treatment regimens that include ineffective medicines, which might lead to emergence of further *M tuberculosis* drug resistance, comprehensive DST results must be available to treating physicians.^2^ This poses enormous challenge to detection, appropriate management and surveillance of XDR-TB in the WHO European region, and even more so in other, resource-poor settings of the world. New genotypic DST solutions, like Deeplex Myc-TB® (Genoscreen, Lille, France), may have the potential to improve rapid access to DST.^27^

All sites in the survey reported availability of treatment for DS-TB. However, as the number of drugs to which the strain is resistant increases, the availability of suitable regimens declined. Middle income countries have generally less resistance-appropriate treatment options then high income countries. In the 2013 TBnet assessment on the availability of regimens performed in 37 European countries, 81% of countries had access to adequate MDR/RR-TB regimens (now 72%), 65% to a pre-XDR TB regimen (now 67%), and 35% to a XDR-TB regimen (now 42%). ^10^ Unfortunately, the access situation to the relevant regimens seems fairly unchanged since the introduction of new drugs and regimens.

The limited availability of medicines for patients with MDR/RR-TB and even more with XDR-TB dramatically affects the quality of care of the 70 000 patients affected by MDR/RR-TB including 7 259 patients affected by XDR-TB notified in the WHO European region in 2019,^28^ whose treatment options remain limited. High cost of novel drugs for anti-tuberculosis treatment regimens is restricting availability of these medicines for many of the affected patients in this region. Based on the findings of this study, the drivers for limited access to TB drugs and DST testing need further investigation and action. This process is already initiated by WHO Regional Office for Europe (T. Humbert, personal communication). An analysis on barriers and challenges to access TB medicines is being conducted, focussing first on EU member states, to be concluded in 2022.

The drug treatment for a patient with MDR/RR-TB with the shorter regimen (including 6 months bedaquiline) costs approximately 18 times more in middle income countries and 106 times more in high income countries than the standardized DS-TB regimen. This has enormous cost implications for health systems in countries with high burden of drug-resistant tuberculosis. For example, the Republic of Moldova (total population of 2·6 million 2020) reported 559 patients with MDR/RR-TB in 2019, corresponding to 73% of all incident MDR/RR-TB patients notified of the whole European Union (763 patients in 28 countries/ total population of 447 million 2021).^28^

The global drug facility provides subsidized access to anti-tuberculosis drugs. Across the WHO European region, 9 countries (including one high income country) report drug procurement via the global drug facility. The shorter MDR/RR-TB regimen with 6 months bedaquiline costs approximately €390 (€1 = USD 0·85) in 2021, using drugs purchased via the global drug facility.^29^ In our study, the median costs for the same regimen were €764 and €29 765 in middle income and high income countries, respectively. The barriers to procurement via global drug facility are not clear and should be investigated and addressed for eligible middle income countries.

Of note, high regimen costs in Europe are related to the high prices of tuberculosis drugs in general, but are impacted in particular by the enormous cost of the new drugs bedaquiline and delamanid. Recent data from Finland and Estonia confirm the very high cost contribution of the new drugs to regimen cost.^7^ In Germany, for instance, the daily therapy cost for bedaquiline is €133. Since July 2020 139 countries, including Eastern European countries, are eligible to procure bedaquiline for €1·27 (€1 = USD 0·85) daily treatment cost via the global drug facility. In contrast, it was estimated that the generic production of bedaquiline could lead to daily treatment cost of €0·21 – 0·48 per day.^30^ Patent protection for bedaquiline lasts at least until June 2023:^29^ only then less expensive generic products could be licensed. New patent applications by the manufacturer of bedaquiline could extent the patent protection till 2027,^29^ despite the fact that the public investments on bedaquiline development exceed the originators company investments by a factor 1·6-5·1.^30^

Rifapentine has recently shown the potential to shorten DS-TB treatment when used in combination with moxifloxacin, isoniazid and pyrazinamide as part of a four-month regimen^31^ which was already endorsed by WHO.^32^ In addition, rifapentine is also recommended for tuberculosis prevention in the one month daily rifapentine/isoniazid (1HP) and three months weekly rifapentine/isoniazid (3HP) regimen.^33^ Of concern, our results show that rifapentine is only available in two middle income countries and four high income countries in the European region. Rifapentine is not registered in the EU. The manufacturer of rifapentine, an off patent protection drug, has withdrawn its patent application with the European Patent Office for a fixed dose combination with isoniazid in 2019.^34^ UNITAID, the Stop TB Partnership and the manufacturer announced in 2019 an access programme for 3HP, which reduced the price of a three-month course of rifapentine to approximately €13·86. Among the countries included in this survey, Russia, the Republic of Moldova and Ukraine were granted such access.^35^ Access to rifapentine in Europe and worldwide remains an urgent matter. This has been addressed very recently with an invitation to manufacturers by the WHO to present rifapentine to the WHO pre-qualification process.^36^

This study has several limitations. Most importantly, it is not fully representative of the access and cost for DST, drugs and regimens in the WHO European region. Data are derived only from a single site in each country, which was considered to represent the access and cost situation in the country. We assumed that data informants were able to gather representative information for their site, which would reflect the country-wide situation. Hence, we refer to countries instead of individual sites within our analysis. Second, data were predominantly obtained from clinicians treating drug-resistant tuberculosis. Data from such a source might differ from data obtained from a national TB programme. Third, we did not analyse the role of possible stock outs on drug availability. Fourth, the choice of regimens for cost calculations followed the recommendations of WHO ^4^ and ATS/CDC/IDSA/ERS ^15^, whereas other regimen compositions could also be possible.

Despite these weaknesses, the study provides first-hand insight about the access to DST, drugs and related drug and regimen costs and should inform health policy decisions in the context of the endTB strategy in Europe.^37^

In conclusion, data provided from this study call for urgent action. Availability of novel drugs and treatment regimens for patients affected by MDR/RR-TB is substantially limited in Europe. Even more limited is the DST capacity for second-line drugs, leading to uncontrolled use of new/re-purposed drugs and the risk of amplifying *M tuberculosis* drug-resistance. Strong political support and coordinated action from civil society, national tuberculosis programmes and non-governmental organizations is needed to ensure access to the best standard of care to patients affected by tuberculosis.

## Data Availability

All data produced in the present study are available upon reasonable request to the authors

## Members of the TBnet study group

### We thank the following colleagues for the contribution of data to this analysis

Hasan Hafizi (Tirana, Albania), Naira Khachatryan, Harut Aroyan, Eduard Kabasakalyan (Yerevan, Armenia), Michael Knappik (Vienna, Austria), Alena Skrahina, Dzmitry Klimuk, Alena Nikolenka (Minsk, Belarus), Inge Muylle (Brussels, Belgium), Vladimir Milanov, Desislava Velkovska, Neli Tarinska, Elizabeta Bachiyska (Sofia, Bulgaria), Mateja Jankovic (Zagreb, Croatia), Despo Pieridou, Tonia Adamide, Nicos Nicolaou (Nicosia, Cyprus), Martina Vasakova, Mariia Sukholytka, Emilia Kopeckà, Beata Polakova (Prague, Czech Republic), Aase Bengard Anderson, Dorte Bek Folkvardsen, Erik Svensson (Copenhagen, Denmark), Manfred Danilovits, Tiina Kummik (Tartu, Estonia), Tuula Vasankari (Turku, Finland), Mathilde Fréchet-Jachym, Audrey Nahmiash (Briis-sous-Forges, France), Tamar Togonidze, Zaza Avaliani, Inga Kinkladze, Rusudan Aspindzelashvili, Teona Bichashvili, Gulnazi Losaberidze, Tsitsino Merabishvili (Tbilisi, Georgia), Barbara Kalsdorf (Borstel, Germany), Katerina Manika, Karyofyllis Tsiakitzis (Thessaloniki, Greece), Agnes Bakos (Budapest, Hungary), Tinna Rán Ægisdóttir, Guðrún Svanhvít Michelsen, Kristín Karlsdóttir (Reykjavik, Iceland), Anne-Marie McLaughlin, Margaret Fitzgibbon (Dublin, Ireland), Daniel Chemtob (Jerusalem, Israel), Luigi R. Codecasa, Maurizio Ferrarese, Stefania Torri (Milano, Italy), Majlinda Gjocaj (Pristina, Kosovo), Liga Kuksa (Riga, Latvia), Edita Davidaviciene (Vilnus, Lithuania), Gil Wirtz, Monique Perrin (Luxembourg, Luxembourg), Analita Pace Asciak (Valetta, Malta), Dumitri Chesov (Chisinau, Moldova), Wiel de Lange, Onno Akkerman (Groningen, Netherlands), Biljana Illievska-Poposka (Skopje, North Macedonia), Ulrich Mack, Mogens Jesenius, Lajla Kvalvik, Anne Torunn Mengshoel (Oslo, Norway), Katarzyna Kruzak (Krakow, Poland), Raquel Duarte, Nadine Ribeiro (Porto, Portugal), Elmira Ibraim (Bucharest, Romania), Anna Kaluzhenina, Olga Barkanova (Volgograd, Russia), Dragica Pesut (Belgrade, Serbia), Ivan Solovic (Vysne Hagy, Slovakia), Petra Svetina (Golnik, Slovenia), Maria-Luiza de Souza-Galvão, Joan-Pau Millet, Xavi Casas, Montserrat Vives (Barcelona, Spain), Judith Bruchfeld, Paulina Dalemo, Jerker Jonsson (Stockholm, Sweden), Katrin Aeschbacher, Peter Keller (Bern, Switzerland), Seref Ozkara (Ankara, Turkey), Simon Tiberi, Christabelle Chen (London, UK), Yana Terleeva (Kiev, Ukraine) Andrii Dudnyk (Vinnytsia, Ukraine)

## Author contributions

LG, GG, CLa and FvL designed the study, GG and CLe collected data, FvL and GG did the analysis, GG, CLa, LG, FvL drafted the manuscript and all authors reviewed and agreed on the final version for submission. Contributors of the TBnet study group approved the manuscript.

## Role of funding sourc

CLa is supported by the German Center for Infection Research (DZIF). All other authors have no funding source in the context of this manuscript.

## Data sharing statement

Data on availability of drugs, DST and cost are available for sharing immediately after publication for researchers with a sound proposal.

## Supplement

**Figure S1:**
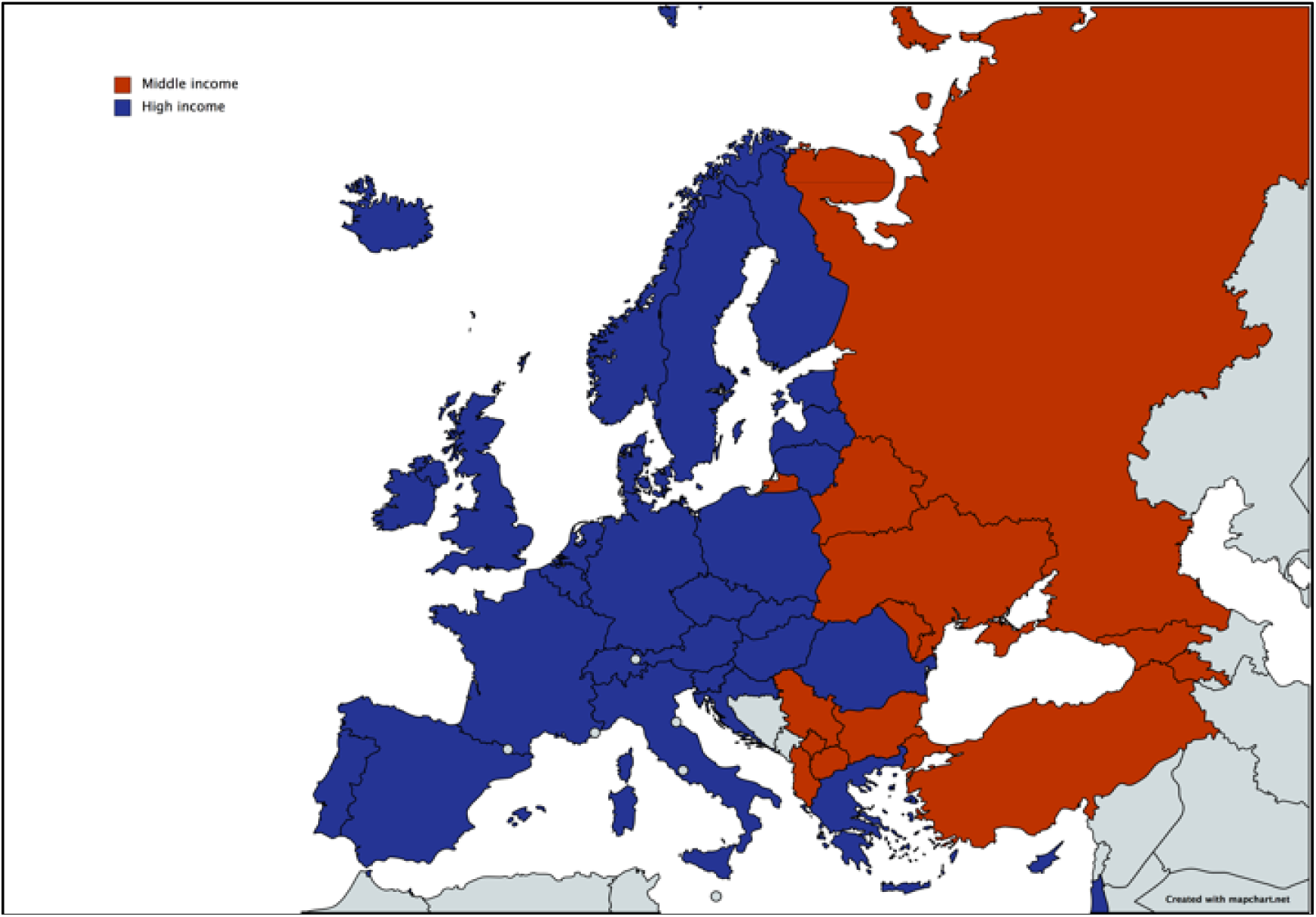
Countries represented in the survey. (high income countries: *Austria, Belgium, Croatia, Czech Republic, Cyprus, Denmark, Estonia, Finland, France, Germany, Greece, Hungary*, Iceland, *Ireland*, Israel, *Italy, Latvia, Lithuania, Luxemburg, Netherlands*, Norway, *Spain, Poland, Portugal, Romania Slovakia, Slovenia, Sweden*, Switzerland, United Kingdom; upper-middle income countries: Albania, Armenia, Belarus, *Bulgaria*, Georgia, Kosovo, North Macedonia, Russia, Serbia, Turkey, lower middle income: Republic of Moldova, Ukraine. In italic: member states of the European Union.)

**Table S1:** Regimen composition and treatment duration (months) by drug according to drug resistance (dosing for a patient with 70 kg body weight).

| Regimen | H | hdH | R | Lfx | Bdq | Lzd | Cfz | Cs | Eto/Pto | Am | Mpm/Amx-Clv | Dlm | Z | E | PAS | Total duration |
| --- | --- | --- | --- | --- | --- | --- | --- | --- | --- | --- | --- | --- | --- | --- | --- | --- |
| DS-TB<sup>†</sup> | 6 |  | 6 |  |  |  |  |  |  |  |  |  | 2 | 2 |  | 6 |
| MDR/RR-TB short, Bdq 6 or 9 months<sup>†</sup> |  | 4 |  | 9 | 6/9* |  | 9 |  | 4 |  |  |  | 9 | 9 |  | 9 |
| MDR/RR-TB long, Bdq 6 or 18 months<sup>†</sup> |  |  |  | 18 | 6/18* | 18 | 18 | 18 |  |  |  |  |  |  |  | 18 |
| pre-XDR TB, using Dlm<sup>†</sup> |  |  |  |  | 20 | 20 | 20 | 20 |  |  |  | 20 |  |  |  | 20 |
| pre-XDR TB, using Am<sup>†</sup> |  |  |  |  | 20 | 20 | 20 | 20 |  | 6 |  |  |  |  |  | 20 |
| XDR-TB, resistant Fq, Bdq, using carbapenem<sup>†</sup> |  |  |  |  |  | 20 | 20 | 20 | 20 |  | 6 | 20 |  |  |  | 20 |
| XDR-TB, resistant Fq, Bdq, using Am |  |  |  |  |  | 20 | 20 | 20 | 20 | 6 |  | 20 |  |  |  | 20 |
| XDR-TB, resistant Fq, Lzd, using carbapenem<sup>†</sup> |  |  |  |  | 20 |  | 20 | 20 | 20 |  | 6 | 20 |  |  |  | 20 |
| XDR-TB, resistant Fq, Lzd, using Am |  |  |  |  | 20 | 20 | 20 | 20 |  | 6 |  | 20 |  |  |  | 20 |
| XDR-TB, resistant Fq, Bdq, Lzd, using carbapenem<sup>†</sup> |  |  |  |  |  |  | 20 | 20 | 20 |  | 6 | 20 |  |  | 20 | 20 |
| XDR-TB, resistant Fq, Bdq, Lzd using Am |  |  |  |  |  |  | 20 | 20 | 20 | 6 |  | 20 |  |  | 20 | 20 |

Abbreviation and dose (once daily if not indicated otherwise), per drug: H - isoniazid 300 mg/d, hdH - isoniazid 600 mg/d; R - rifampicin 600 mg/d; Lfx - levofloxacin 1000 mg/d; Bdq - bedaquiline 200 mg thrice weekly; Lzd - linezolid 600 mg/d, Cfz - clofazimine 100 mg/d; Cs - cycloserine 750 mg/d, Eto/Pto - ethionamide/prothionamide 1000 mg/d; Am - amikacin 1000 mg/d; Mpm/Amx-Clv - meropenem 3 g/d plus amoxicillin/clavulanic acid 3.6 g/d; Dlm - delamanid 200 mg/d; Z - pyrazinamide 2000 mg/d, E - ethambutol 1200 mg/d; PAS - para-amino salicylic-acid 12 g/d.
\*indicates alternatives in duration of treatment with bedaquiline; <sup>†</sup>indicates regimen presented in the main paper, cost for remaining regimens are in supplement; DS-TB – drug susceptible tuberculosis, MDR/RR-TB – rifampicin-resistant/multidrug-resistant tuberculosis, pre-XDR-TB – pre-extensively drug-resistant tuberculosis, XDR-TB – extensively drug-resistant tuberculosis, Fq - fluoroquinolones (moxifloxacin or levofloxacin), carbapenem (meropenem or imipenem).

**Table S2:** Availability and cost of additional regimens for MDR/RR-TB and XDR-TB treatment in countries the WHO Europe region, stratified by world bank income classification^*^, in Euros.

|  | Middle income countries |  |  |  | High income countries |  |  |  |
| --- | --- | --- | --- | --- | --- | --- | --- | --- |
|  | Availability | Cost |  |  | Availability | Cost |  |  |
| Regimen | N (%) | median | min | max | N (%) | median | min | max |
| MDR/RR-TB short, Bdq 9 months<sup>†</sup> | 6 (50.0) | 815 | 582 | 17 040 | 20 (64.5) | 33 087 | 12 422 | 45 035 |
| MDR/RR-TB long, Bdq 6 months<sup>†</sup> | 7 (58.3) | 2 214 | 962 | 16 255 | 24 (77.4) | 51 671 | 16 284 | 178 588 |
| XDR-TB, resistant Fq, Bdq, using Am | 4 (33.3) | 6 424 | 6 233 | 8 221 | 14 (45.2) | 128 873 | 34 405 | 262 804 |
| XDR-TB, resistant Fq, Lzd using Am | 4 (33.3) | 7 216 | 7 175 | 8 758 | 13 (41.9) | 203 644 | 62 955 | 327 399 |
| XDR-TB, resistant Fq, Bdq, Lzd using Am | 4 (33.3) | 6 832 | 6 206 | 8 527 | 11 (35.5) | 147 035 | 43 596 | 278 597 |

\*drug availability data are from 43 countries; regimen cost calculation is based on data from 41 countries, not including Malta and Israel (both high income); <sup>†</sup>refers to the length of bedaquiline treatment; Fq - fluoroquinolone, Bdq - bedaquiline, Am - amikacin, Lzd - linezolid, MDR/RR-TB - multidrug-resistant/rifampicin-resistant tuberculosis, XDR-TB - extensively drug-resistant tuberculosis.

**Table S3:** Cost of regimen for susceptible TB, MDR/RR-TB, pre-XDR TB and XDR-TB in countries in the WHO Europe region, stratified by world bank income classification^†^, in ID$.

|  | Middle income countries |  |  | High income countries |  |  |
| --- | --- | --- | --- | --- | --- | --- |
|  | Cost |  |  | Cost |  |  |
| Regimen* | median | min | max | median | min | max |
| DS-TB | 141 | 68 | 504 | 371 | 145 | 1 279 |
| MDR/RR-TB short, Bdq for 6 months<sup>‡</sup> | 2 891 | 1 984 | 42 785 | 39 644 | 21 347 | 60 616 |
| MDR/RR-TB short, Bdq for 9 months<sup>‡</sup> | 3 088 | 2 129 | 48 118 | 44 366 | 23 709 | 67 547 |
| MDR/RR-TB long, Bdq for 6 months<sup>‡</sup> | 7 616 | 4 081 | 45 903 | 70 821 | 29 903 | 210 664 |
| MDR/RR-TB long, Bdq for 18 months<sup>‡</sup> | 10 162 | 6 094 | 119 949 | 128 103 | 62 697 | 255 498 |
| pre-XDR TB, using Dlm | 31 348 | 25 597 | 45 128 | 235 913 | 117 503 | 438 633 |
| pre-XDR TB, using Am | 10 023 | 7 343 | 11 343 | 146 910 | 68 703 | 294 385 |
| XDR-TB, resistant Fq Bdq, using Carbapenem<sup>#</sup> | 32 926 | 29 656 | 38 597 | 160 755 | 64 211 | 301 450 |
| XDR-TB, resistant Fq, Bdq, using Am | 27 912 | 23 557 | 28 924 | 152 969 | 58 149 | 310 007 |
| XDR-TB, resistant Fq Lzd, using Carbapenem<sup>#</sup> | 36 831 | 32 447 | 40 444 | 260 272 | 167 416 | 377 648 |
| XDR-TB, resistant Fq Lzd, using Am | 30 478 | 26 378 | 33 516 | 248 325 | 115 609 | 386 204 |
| XDR-TB, resistant Fq Bdq, Lzd, using Carbapenem<sup>#</sup> | 33 496 | 31 501 | 39 651 | 175 027 | 155 533 | 320 080 |
| XDR-TB, resistant Fq Bdq, Lzd using Am | 28 839 | 25 402 | 29 330 | 162 600 | 80 059 | 328 637 |

\* detailed regimen composition depicted in table S1; <sup>†</sup> drug cost were obtained in 41 countries, not including Malta and Israel; <sup>‡</sup> refers to the length of bedaquiline (Bdq) treatment; <sup>#</sup> Carbapenem, refers to use of cheapest available carbapenem (Meropenem or Imipenem); MDR/RR-TB – multidrug-resistant/rifampicin-resistant tuberculosis, pre-XDR TB – pre-extensively drug-resistant tuberculosis, XDR-TB – extensively drug-resistant tuberculosis, Fq - fluoroquinolone, Dlm - delamanid, Am - amikacin, Lzd - linezolid.

**Table S4:** Cost of individual TB drugs per treatment day for a model patient with 70kg body weight, in countries in the WHO Europeregion^*†^, in Euros.

| Drug name | Middle income countries |  |  |  |  | High income countries |  |  |  |  |
| --- | --- | --- | --- | --- | --- | --- | --- | --- | --- | --- |
|  | Drug available, N of countries | Cost data available, N of countries | Cost of individual drug for daily treatment of a 70 kg patient |  |  | Drug available, N of countries | Cost data available, N of countries | Cost of individual drug for daily treatment of a 70 kg patient |  |  |
|  |  |  | median | min | max |  |  | median | min | max |
| **First line** |  |  |  |  |  |  |  |  |  |  |
| Rifampicin<sup>**</sup> | 10 | 10 | 0.19 | 0.03 | 0.41 | 31 | 29 | 0.65 | 0.18 | 2.94 |
| Isoniazid | 12 | 12 | 0.05 | 0.01 | 0.12 | 31 | 29 | 0.80 | 0.08 | 11.07 |
| Pyrazinamide | 12 | 12 | 0.12 | 0.04 | 0.84 | 31 | 28 | 0.76 | 0.16 | 2.80 |
| Ethambutol | 12 | 12 | 0.15 | 0.06 | 0.48 | 31 | 29 | 0.72 | 0.09 | 4.71 |
| **Group A** |  |  |  |  |  |  |  |  |  |  |
| Bedaquiline<sup>‡</sup> | 9 | 9 | 1.60 | 1.30 | 62.26 | 27 | 25 | 103.98 | 36.55 | 150.95 |
| Linezolid | 12 | 12 | 1.85 | 0.16 | 25.65 | 31 | 29 | 17.03 | 1.00 | 91.43 |
| Moxifloxacin | 11 | 11 | 0.21 | 0.08 | 2.33 | 30 | 28 | 1.59 | 0.39 | 11.54 |
| Levofloxacin | 12 | 12 | 0.14 | 0.01 | 1.44 | 31 | 29 | 2.28 | 0.34 | 108.86 |
| **Group B** |  |  |  |  |  |  |  |  |  |  |
| Terizidone<sup>#</sup> | 2 | 2 | 4.74 | 3.59 | 5.88 | 9 | 9 | 39.60 | 18.00 | 67.98 |
| Cycloserine<sup>#</sup> | 11 | 11 | 0.66 | 0.48 | 6.81 | 25 | 23 | 15.90 | 2.07 | 280.14 |
| Clofazimine | 8 | 8 | 0.75 | 0.41 | 11.10 | 27 | 25 | 2.23 | 0.15 | 3.76 |
| **Group C** |  |  |  |  |  |  |  |  |  |  |
| Delamanid | 6 | 6 | 8.52 | 8.04 | 83.48 | 18 | 16 | 128.04 | 12.92 | 190.56 |
| Meropenem | 3 | 3 | 8.88 | 5.79 | 16.68 | 25 | 24 | 26.91 | 7.77 | 126.09 |
| Imipenem | 4 | 4 | 14.64 | 10.28 | 17.36 | 21 | 19 | 23.40 | 8.28 | 127.48 |
| Amoxicillin/Clavulanic acid | 7 | 7 | 0.20 | 0.15 | 0.81 | 29 | 26 | 0.63 | 0.15 | 5.88 |
| Streptomycin | 2 | 2 | 0.20 | 0.02 | 0.39 | 15 | 11 | 6.50 | 0.90 | 76.79 |
| Amikacin | 7 | 7 | 1.18 | 0.26 | 2.76 | 30 | 28 | 10.10 | 2.42 | 246.76 |
| Ethionamide<sup>§</sup> | 3 | 3 | 0.40 | 0.32 | 0.80 | 13 | 10 | 28.36 | 4.40 | 33.48 |
| Prothionamide<sup>§</sup> | 8 | 8 | 0.36 | 0.28 | 1.84 | 14 | 14 | 5.36 | 0.76 | 9.68 |
| PAS acid | 8 | 5 | 2.46 | 0.24 | 3.72 | 15 | 15 | 49.17 | 24.81 | 74.79 |
| PAS salt | 8 | 5 | 3.78 | 2.46 | 17.94 | 2 | 2 | 46.02 | 27.81 | 64.23 |
| **Other** |  |  |  |  |  |  |  |  |  |  |
| Rifabutin | 2 | 1 | 1.56 | 1.56 | 1.56 | 25 | 22 | 7.40 | 3.18 | 23.56 |
| Rifapentine | 2 | 2 | 3.18 | 3.12 | 3.24 | 4 | 3 | 52.26 | 4.80 | 53.10 |
| Pretomanid | 0 | 0 |  |  |  | 4 | 3 | 33.02 | 27.90 | 44.62 |
| Capreomycin | 6 | 6 | 2.77 | 1.64 | 3.08 | 14 | 11 | 43.77 | 3.22 | 97.00 |
| Kanamycin | 5 | 5 | 2.16 | 0.52 | 4.00 | 5 | 3 | 31.80 | 5.56 | 31.80 |

\* drug availability data are from 43 countries, drug cost calculation is based on data from 41 countries, not including Malta and Israel; † daily drug dosage used for calculation of treatment cost: isoniazid 300 mg/d; rifampicin 600 mg/d; pyrazinamide 2000 mg/d; ethambutol 1200 mg/d; bedaquiline 200 mg 3x week; linezolid 600 mg/d; levofloxacin 1000 mg/d; moxifloxacin 400 mg/d; clofazimine 100 mg/d; cycloserine/terizidone 750 mg/d; delamanid 200 mg/d; imipenem 2000 mg/ d; meropenem 3000 mg/d; amoxicillin/clavulanic acid 1500/375 mg/d or 3000/600 mg (depending on availability); streptomycin 1000 mg/d; amikacin 1000 mg/d; ethionamide/prothionamide 1000 mg/d; PAS acid /PAS sodium salt 12 g/d; rifabutin 300 mg/d; rifapentine 900
mg/d; pretomanid 200 mg/d; capreomycin 1000 mg/d; kanamycin 1000 mg/d; <sup>‡</sup>bedaquiline 200 x 3 weekly: daily cost calculated as (200 mg x 3)/7 to estimate daily treatment cost; <sup>#</sup>cycloserine or terizidone are available in 11 (25.6%) middle income countries and 30 (69.7%) high income countries; <sup>§</sup>protionamide or ethionamide are available in 9 (20.9%) middle income countries and 24 (55.8%) high income countries; <sup>\*\*</sup>rifampicin was available only as fixed dose combination in Albania and Kosovo.

## References

1. Dheda K, Gumbo T, Maartens G, et al. The Lancet Respiratory Medicine Commission: 2019 update: epidemiology, pathogenesis, transmission, diagnosis, and management of multidrug-resistant and incurable tuberculosis. The Lancet Respiratory medicine 2019; 7(9): 820–6.

2. Lange C, Dheda K, Chesov D, Mandalakas AM, Udwadia Z, Horsburgh CR, Jr. Management of drug-resistant tuberculosis. Lancet 2019; 394(10202): 953–66.

3. World Health Organization. Global tuberculosis report 2020, Licence: CC BY-NC-SA 3.0 IGO., 2020.

4. World Health Organization. WHO consolidated guidelines on tuberculosis. Module 4: treatment - drug-resistant tuberculosis treatment. World Health Organization; 2020. Licence: CC BY-NC-SA 3.0 IGO., 2020.

5. MSF Access Campaign. DR-TB and TB prevention drugs under the microscope, 2020. https://msfaccess.org/dr-tb-and-tb-prevention-drugs-under-microscope-7th-edition (accessed 07.10.2021)

6. World Health Organization. WHO operational handbook on tuberculosis. Module 4: treatment - drug-resistant tuberculosis treatment. Geneva 2020. Licence: CC BY-NC-SA 3.0 IGO., 2020.

7. Feuth T, Patovirta RL, Grierson S, et al. Costs of multidrug-resistant TB treatment in Finland and Estonia affected by the 2019 WHO guidelines. Int J Tuberc Lung Dis 2021; 25(7): 554–9.

8. Manalan K, Green N, Arnold A, et al. A cost comparison of amikacin therapy with bedaquiline, for drug-resistant tuberculosis in the UK. J Infect 2020; 80(1): 38–41.

9. van Leth F, Brinkmann F, Cirillo DM, et al. The Tuberculosis Network European Trials group (TBnet) ERS Clinical Research Collaboration: addressing drug-resistant tuberculosis through European cooperation. Eur Respir J 2019; 53(1):1802089.

10. Günther G, Gomez GB, Lange C, Rupert S, van Leth F, Tbnet. Availability, price and affordability of anti-tuberculosis drugs in Europe: a TBNET survey. Eur Respir J 2015; 45(4): 1081–8.

11. Global Drug Facility. Medicine Catalogue 2020. https://www.stoptb.org/sites/default/files/gdfmedicinescatalog_1.pdf (accessed 20.10.2021).

12. World Bank. Purchasing Power Parities and the Size of World Economies: Results from the 2017 International Comparison Program. Washington, DC: World Bank. DOI: 10.1596/978-1-4648-1530-0. License: Creative Commons Attribution CC BY 3.0 IGO, 2017.

13. World Bank. World Bank Country and Lending Groups 2021. https://datahelpdesk.worldbank.org/knowledgebase/articles/906519-world-bank-country-and-lending-groups (accessed 13.08.2021).

14. World Health Organization. Guidelines for treatment of drug-susceptible tuberculosis and patient care, 2017 update. Geneva: World Health Organization; 2017. Licence: CC BY-NC-SA 3.0 IGO., 2017.

15. Nahid P, Mase SR, Migliori GB, et al. Treatment of Drug-Resistant Tuberculosis. An Official ATS/CDC/ERS/IDSA Clinical Practice Guideline. Am J Respir Crit Care Med 2019; 200(10): e93–e142.

16. Viney K, Linh NN, Gegia M, et al. New definitions of pre-extensively and extensively drug-resistant tuberculosis: update from the World Health Organization. Eur Respir J 2021; 57(4):2100361.

17. Conradie F, Diacon AH, Ngubane N, et al. Treatment of Highly Drug-Resistant Pulmonary Tuberculosis. N Engl J Med 2020; 382(10): 893–902.

18. Farooq HZ, Cirillo DM, Hillemann D, et al. Limited Capability for Testing Mycobacterium tuberculosis for Susceptibility to New Drugs. Emerg Inf Dis 2021; 27(3): 985–7.

19. Köser CU, Maurer FP, Kranzer K. ‘Those who cannot remember the past are condemned to repeat it’: Drug-susceptibility testing for bedaquiline and delamanid. Int J Infect Dis 2019; 80S: S32–S5.

20. He W, Liu C, Liu D, et al. Prevalence of Mycobacterium tuberculosis resistant to Bedaquiline and Delamanid in China. J Glob Antimicrob Resist 2021; 26:241–248.

21. Lee T, Lee SJ, Jeon D, et al. Additional Drug Resistance in Patients with Multidrug-resistant Tuberculosis in Korea: a Multicenter Study from 2010 to 2019. J Korean Med Sci 2021; 36(26): e174.

22. Nimmo C, Millard J, van Dorp L, et al. Population-level emergence of bedaquiline and clofazimine resistance-associated variants among patients with drug-resistant tuberculosis in southern Africa: a phenotypic and phylogenetic analysis. Lancet Microbe 2020; 1(4): e165–e74.

23. Roelens M, Battista Migliori G, Rozanova L, et al. Evidence-based Definition for Extensively Drug-Resistant Tuberculosis. Am J Resp Crit Care Med 2021; 204(6): 713–22.

24. Ismail NA, Omar SV, Moultrie H, et al. Assessment of epidemiological and genetic characteristics and clinical outcomes of resistance to bedaquiline in patients treated for rifampicin-resistant tuberculosis: a cross-sectional and longitudinal study. Lancet Inf Dis 2021; S1473-3099(21)00470-9.

25. Alagna R, Cabibbe AM, Miotto P, et al. Is the new WHO definition of extensively drug-resistant tuberculosis easy to apply in practiceã Eur Respir J 2021; 58(1):2100959.

26. Veziris N, Bonnet I, Morel F, et al. Impact of the revised definition of extensively drug resistant tuberculosis. Eur Respir J 2021; 58(2): 2100641.

27. Jouet A, Gaudin C, Badalato N, et al. Deep amplicon sequencing for culture-free prediction of susceptibility or resistance to 13 anti-tuberculous drugs. Eur Respir J 2021; 57(3): 2002338.

28. WHO/ECDC. Tuberculosis Surveillance and monitoring in Europe 2021 -2019 data. 2021. https://www.ecdc.europa.eu/en/publications-data/tuberculosis-surveillance-and-monitoring-europe-2021-2019-data(accessed 07.10.2021).

29. MSF Access Campaign. DR-TB and TB prevention drugs under the microscope, 2019. https://msfaccess.org/dr-tb-and-tb-prevention-drugs-under-microscope-6th-edition (accessed 07.10.2021).

30. Gotham D, Fortunak J, Pozniak A, et al. Estimated generic prices for novel treatments for drug-resistant tuberculosis. Journal Antimicrob Chemother 2017; 72(4): 1243–52.

31. Dorman SE, Nahid P, Kurbatova EV, et al. Four-Month Rifapentine Regimens with or without Moxifloxacin for Tuberculosis. N Engl J Med 2021; 384(18): 1705–18.

32. World Health Organization. Treatment of drug-susceptible tuberculosis - rapid communication. 2021. https://www.who.int/publications/i/item/9789240028678 (accessed 07.10.2021).

33. World Health Organization. WHO consolidated guidelines on tuberculosis. Module 1: prevention – tuberculosis preventive treatment. Geneva; 2020. Licence: CC BY-NC-SA 3.0 IGO.

34. Treatment Action Group. Isoniazid/Rifapentine (3HP) Access Roadmap and Patent Landscape. 2020. https://www.treatmentactiongroup.org/wp-content/uploads/2020/03/3hp_access_roadmap_and_patent_landscape.pdf (accessed 07.10.2021).

35. Stop TB Partnership and The Global Fund. Rifapentine Global Price Discount. 2019. https://www.impaact4tb.org/wpcontent/uploads/2019/11/Rifapentine_Global_Price_Discount_Communique.pdf (accessed 07.10.2021).

36. World Health Organization. New Invitation for Expression of Interest (EOI) to manufacturers of antituberculosis medicines published. 2021. https://extranet.who.int/pqweb/news/new-invitation-expression-interest-eoi-manufacturers-antituberculosis-medicines-published (accessed 20.07.2021).

37. World Health Organization. The WHO End TB Strategy, 2015. https://www.who.int/tb/strategy/End_TB_Strategy.pdf (accessed 20.11.2021).

